# Patient and programmatic characteristics, retention and predictors of attrition among patients starting antiretroviral therapy (ART) before and after the implementation of HIV “Treat All” in Zimbabwe

**DOI:** 10.1101/2020.07.12.20151845

**Authors:** Richard Makurumidze, Jozefien Buyze, Tom Decroo, Lutgarde Lynen, Madelon de Rooij, Trevor Mataranyika, Ngwarai Sithole, Kudakwashe C. Takarinda, Tsitsi Apollo, James Hakim, Wim Van Damme, Simbarashe Rusakaniko

## Abstract

**Background:** Since scale-up of the HIV “Treat All”, evidence on its real-world effect on known predictors of attrition (either death or lost to follow-up) is lacking. We conducted a retrospective study using Zimbabwe ART program data to assess the association between “Treat All” and, patient and programmatic characteristics, retention and predictors of attrition.

**Methods:** We used patient-level data from the electronic patient monitoring system (ePMS) from the nine districts which piloted “Treat All”. We compared patient and programme characteristics, retention and predictors of attrition (lost to follow-up, death or stopping ART) in two cohorts; before (April/May 2016) and after (January/February 2017) “Treat All”. Retention was estimated using survival analysis. Predictors of attrition were determined using a multivariable Cox regression model. Interactions were used to assess the change in predictors.

**Results:** We analysed 3787 patients, 1738 (45.9%) and 2049 (54.1%) started ART before and after “Treat All”, respectively. The proportion of men was higher after “Treat All” (39.4.% vs 36.2%, p=0.044). Same-day ART initiation was more frequent after “Treat All” (43.2% vs 16.4%; p<0.001) than before. Retention on ART was higher before “Treat All” (p<0.001). Among non-pregnant women and men, the adjusted hazard ratio (aHR) of attrition after “Treat All” compared to before “Treat All” was 1.73 (95%CI: 1.30 - 2.31). The observed hazard of attrition for women being pregnant at ART initiation decreased by 17% (aHR: 1.73*0.48 = 0.83). Being male (vs female; aHR: 1.45; 95%CI: 1.12 - 1.87) and WHO Stage IV (vs WHO Stage I-III; aHR: 2.89; 95%CI: 1.16 - 7.11) predicted attrition both before and after “Treat All” implementation.

**Conclusion:** Attrition was higher after “Treat All”; being male, WHO Stage 4, and pregnancy predicted attrition in both before and after Treat All. However, pregnancy became a less strong risk factor for attrition after “Treat All” implementation.”

## Introduction

The highest number of people living with HIV (PLHIV) originate from the East and Southern African (ESA) region. However, this region has not yet met the Joint United Nations Programme for HIV/AIDS (UNAIDS) 90-90-90 targets, which were launched in 2014 to achieve epidemic control by 2020 [1]. By the end of 2018 in the ESA region, 85% [95% confidence interval (CI); 75-95%] of PLHIV knew their HIV status, with 67% [95%CI; 52 -78%] of those diagnosed on antiretroviral therapy (ART) and only 58% [95%CI; 50 -66%] of those on ART achieving sustained viral load suppression [2]. To close the gap, by the end of 2018, 93% of low- and middle-income countries and 100% of those designated as Fast Track countries had adopted the HIV “Treat All” policy [3].

Several clinical trials have shown the positive effects of “Treat All” on ART initiation, linkage to care, virologic suppression, and retention [4,5]. Multiple studies assessed the real-world effects of “Treat All” on ART initiation, linkage, virologic suppression, and retention. However, these studies have reported inconsistent results [6–13]. None of the previous studies assessed if predictors for attrition changed since the transition to “Treat All”. There is also still lack of evidence on the effect of “Treat All” on the patient mix, i.e. change in the uptake of ART services among previously underserved groups outside of trial settings. In Zimbabwe, as in many other African countries, men and adolescents and young adults remain underserved [14–17]. Whether since the scale-up of “Treat All” the uptake of HIV care services, have increased in favour of these previously underserved subgroups is unknown.

Moreover, it is not fully understood how programmatic procedures related to the provision of HIV care across the cascade have been adapted and influence the outcomes.

Since July 2016 Zimbabwe started “Treat All” implementation. We used program data from those districts that piloted “Treat All” before it was expanded to the rest of Zimbabwe. We assessed the association between “Treat All” and, patient and programme characteristics, retention and predictors of attrition by comparing the ART cohorts enrolled during two periods before and after implementation of the “Treat All” policy. This is the first study to assess the performance of the ART programme in Zimbabwe since the start of HIV “Treat All” policy implementation and a follow-up on previous national studies on ART outcomes in the country.

## Methods and Materials

### Study design

A retrospective cohort study using routinely collected individual patient data was conducted.

### Study setting

Zimbabwe has a generalised HIV epidemic with an estimated number of PLHIV of 1.4 million [18]. The country has made significant progress towards achieving the UNAIDS 90-90-90 targets: 76.8% of PLHIV are aware of their HIV status; of that 88.4 % were on ART and among those on ART, 85.3% were virally suppressed [15]. The study used data from the nine districts (Chipinge, Bulilima, Gwanda, Harare, Mangwe, Makoni, Mazowe, Mutasa and Mutare) that piloted “Treat All”. Zimbabwe is geographically divided into ten provinces and 63 districts. The nine pilot districts were purposively selected from 4 provinces supported by the United States Presidential Emergency Plan for AIDS Relief (PEPFAR).

### Implementation of “Treat All” in the pilot districts

The pilot started in July 2016. By August of the same year, all nine pilot districts were implementing “Treat All”. Before the start of “Treat All”, health care workers were trained on the rationale of Treat All, clinical assessment of opportunistic infections, ART initiation, adherence counselling and patient follow-up, tracking and call back of clients who were not ART because of the previous eligibility criteria and quantification of ART commodities. One-day meetings were conducted in all districts to sensitise community stakeholders, including traditional and political leaders, community-based organisations and networks of PLHIV on the new “Treat All” criteria and its benefits. Four technical working groups were established to monitor progress on four different themes: training and capacity building, linkage and referral of clients, learning and documentation and communication. PEPFAR funded the pilot through its two main implementing partners (International Training Education and Centre for Health - I-TECH and Organisation for Public Health Intervention Development - OPHID). The Ministry of Health and Child Care provided the necessary comprehensive support during all stages of the pilot.

### Study population

Health facilities were included if they used an HIV programme electronic medical record, i.e. the electronic patient monitoring system (ePMS), which was mainly used in high-volume sites. Of 385 health facilities in the nine pilot districts, 131 (30%) had ePMS. Of 131 health facilities with ePMS, 72 had updated data in the national database at the end of December 2018 and were included in the study. The before “Treat All” cohort and the after “Treat All” cohort included all patients who started ART between April-May 2016 and January -February 2017, respectively.

### Data sources and study procedures

The patient’s medical data were first captured in a paper-based medical record (Patient ART/Opportunistic Infection Booklet). The data were then entered in the ePMS by either health care workers or data entry clerks. Every month health facilities submit an encrypted back-up of the ePMS data to the district where all data are consolidated. The consolidated district-level data is then submitted to the national level, where it is further consolidated into one national database. Data for this study were extracted from the national database and cleaned (synchronised and deduplicated) before analysis (S1 Figure).

### Study variables

The variables extracted for analysis included patient and programmatic characteristics. Patient characteristics included demographics (age and sex), and baseline clinical parameters at ART initiation (functional status, WHO Stage, pregnancy, tuberculosis status). Programmatic characteristics included HIV testing modality, level of care, ART regimen and time from HIV testing to ART initiation. The patient follow-up status (active on treatment, lost to follow-up (LTFU), dead, transferred or stopped ART) and date were also collected.

### Data analysis

The data were analysed using Stata version 16.0 (Stata Corp, College Station, Texas, USA) [19]. Descriptive statistics (frequencies and proportions or medians and quartiles) were used to describe and compare patient and programmatic characteristics (before and after “Treat All”). The Chi-square or Fisher’s exact test were used to compare the distribution for the categorical patient and programmatic characteristics before “Treat All” with the distribution after “Treat All”. The Wilcoxon rank-sum test and t-test were used for skewed and symmetrical continuous variables, respectively. The adverse outcome event was attrition, which included those who died, those LTFU and those who stopped ART. LTFU were those whose last recorded clinic visit date, or pharmacy pill pick-up date, was ≥180 days before the date of data extraction from the ePMS. Patients were considered active on ART when their last recorded clinic visit date or pharmacy pill pick-up date was <180 days before the date of data extraction from the ePMS. We calculated the proportion of LTFU patients who only visited the health facility once on the day of ART initiation by dividing the number LTFU patients who only visited the health facility once on the day of ART initiation with the total number initiated on ART in the cohort. In the time-to-event analysis, patients classified as active on ART were censored on the date of their most recently recorded clinic visit or pharmacy pick-up while those who transferred out were censored on the date of transfer out. The survival time was calculated as the time from the date of ART initiation to the date of censoring or attrition. Kaplan Meier survival curves and statistics were used to estimate retention at 6, 12 and 24 months. The log-rank test was used to compare survival curves for different strata.

In the bivariate and multivariate analysis, we included variables with less than 30% missingness. Missing data of included variables were managed through imputation. The data were assumed to be missing at random. Multiple imputation was conducted using multiple chains equations, and twenty imputed datasets were created and used for analysis. Cox proportional hazard models with a frailty variable for health facility were used to identify predictors of attrition. Proportional hazard assumptions were tested by comparing observed with predicted survival curves and log-log plots. The cohort variable (before “Treat All”, after “Treat All”) was included in all bivariate and multivariate models. A hierarchical approach was employed. All variables associated with P-value <0.1 in the bivariate analysis were included in the multivariate model. Sex was maintained in the model because of clinical and programmatic relevance. Stepwise backward elimination was used until all variables in the model had a p-value < 0.05. To be able to evaluate whether the hazard of significant predictors changed between the before and after “Treat All” cohort we looked at the interactions between the significant variables and our primary exposure variable (i.e. cohort). They were assessed and added to the model, using the same hierarchical approach and stepwise backward elimination approach described above.

### Ethical (and regulatory) review

This study was submitted for ethical review and approval to the Institutional Review Board (IRB-1257/18) of the Institute of Tropical Medicine, Antwerp, Belgium; University of Zimbabwe Joint Research Ethics Committee (JREC/239/18) and Medical Research Council of Zimbabwe (MRCZ/A/2410). In addition, permission to conduct the study was sought from the Ministry of Health and Child Care. The data were anonymised to maintain privacy and confidentiality.

## Results

### a) Study participants

We analysed 3787 patients; 1738 (45.9%) from the before and 2049 (54.1%) from the after “Treat All” cohorts.

### b) Association between “Treat All” and patient and programmatic characteristics

In the before and after “Treat All” cohort median age was 37 [interquartile range (IQR), 30-44] and 36 (IQR, 29-43) years, respectively. The proportion of men among those who started ART was significantly higher after “Treat All” compared to before “Treat All” cohort (39.4% vs 36.2%, p =0.044). There was no significant increase in the proportion of adolescents among those who started ART. The proportion of asymptomatic patients (WHO Stage 1) was higher in the after “Treat All” cohort (42.5% vs 26.1%, p <0.001) (Table 1).

**Table 1:**
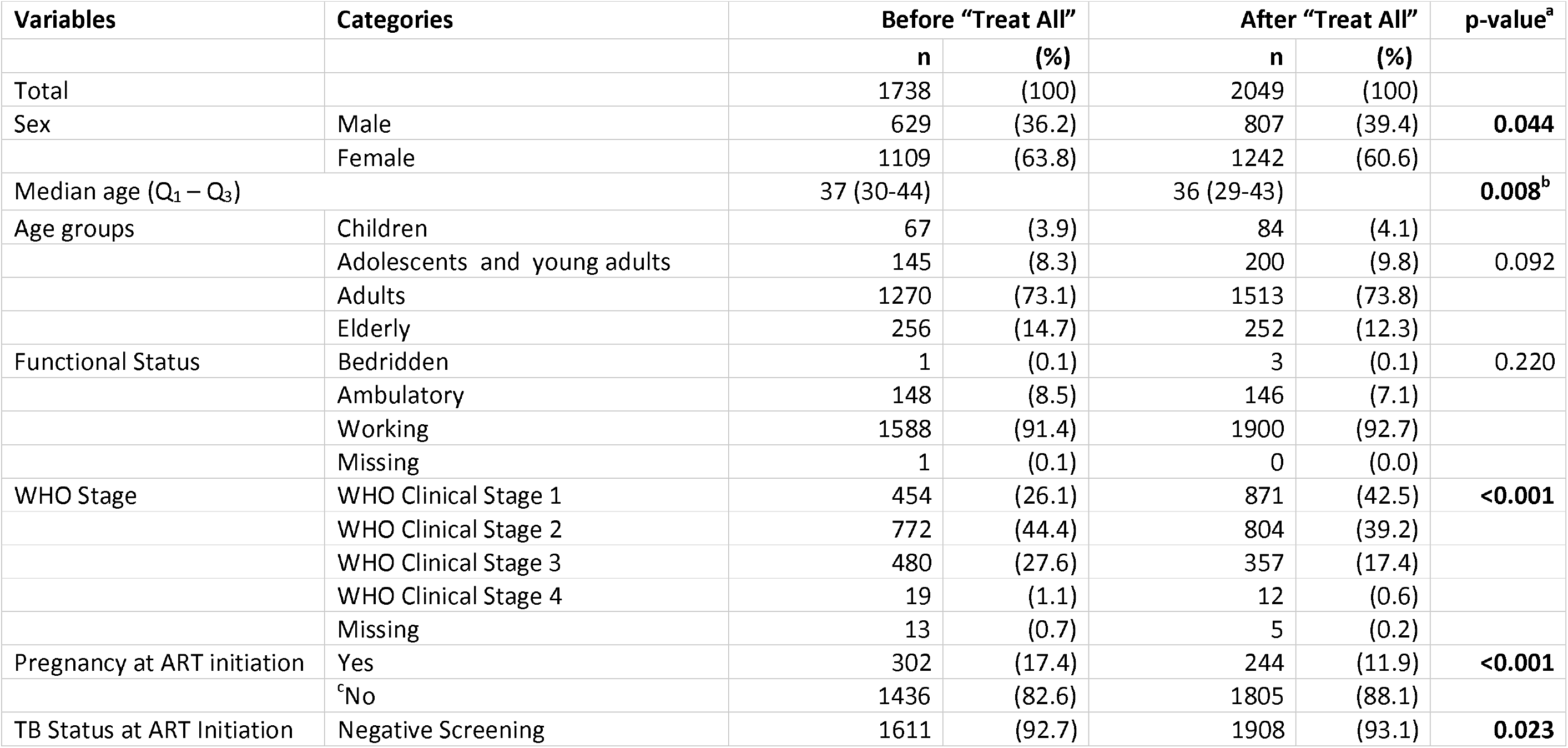
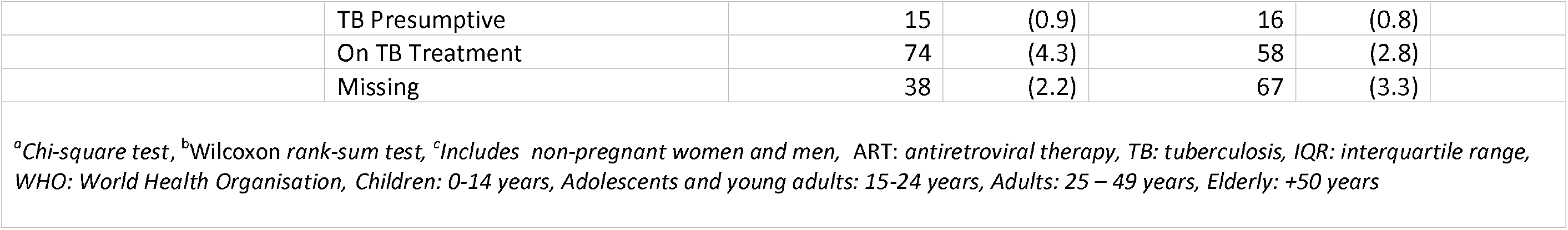
Comparison of baseline patient characteristics starting ART before (April-May 2016) and after (January-February 2017) the implementation of HIV “Treat All” in 9 pilots districts in Zimbabwe.

| Variables | Categories | Before “Treat All” |  | After “Treat All” |  | p-value<sup>a</sup> |
| --- | --- | --- | --- | --- | --- | --- |
|  |  | n | (%) | n | (%) |  |
| Total |  | 1738 | (100) | 2049 | (100) |  |
| Sex | Male | 629 | (36.2) | 807 | (39.4) | **0.044** |
|  | Female | 1109 | (63.8) | 1242 | (60.6) |  |
| Median age (Q<sub>1</sub> – Q<sub>3</sub>) |  | 37 (30-44) |  | 36 (29-43) |  | **0.008<sup>b</sup>** |
| Age groups | Children | 67 | (3.9) | 84 | (4.1) |  |
|  | Adolescents and young adults | 145 | (8.3) | 200 | (9.8) | 0.092 |
|  | Adults | 1270 | (73.1) | 1513 | (73.8) |  |
|  | Elderly | 256 | (14.7) | 252 | (12.3) |  |
| Functional Status | Bedridden | 1 | (0.1) | 3 | (0.1) | 0.220 |
|  | Ambulatory | 148 | (8.5) | 146 | (7.1) |  |
|  | Working | 1588 | (91.4) | 1900 | (92.7) |  |
|  | Missing | 1 | (0.1) | 0 | (0.0) |  |
| WHO Stage | WHO Clinical Stage 1 | 454 | (26.1) | 871 | (42.5) | **<0.001** |
|  | WHO Clinical Stage 2 | 772 | (44.4) | 804 | (39.2) |  |
|  | WHO Clinical Stage 3 | 480 | (27.6) | 357 | (17.4) |  |
|  | WHO Clinical Stage 4 | 19 | (1.1) | 12 | (0.6) |  |
|  | Missing | 13 | (0.7) | 5 | (0.2) |  |
| Pregnancy at ART initiation | Yes | 302 | (17.4) | 244 | (11.9) | **<0.001** |
|  | <sup>c</sup>No | 1436 | (82.6) | 1805 | (88.1) |  |
| TB Status at ART Initiation | Negative Screening | 1611 | (92.7) | 1908 | (93.1) | **0.023** |
|  | TB Presumptive | 15 | (0.9) | 16 | (0.8) |  |
|  | On TB Treatment | 74 | (4.3) | 58 | (2.8) |  |
|  | Missing | 38 | (2.2) | 67 | (3.3) |  |

<sup>a</sup>Chi-square test, <sup>b</sup>Wilcoxon rank-sum test, <sup>c</sup>Includes non-pregnant women and men, ART: antiretroviral therapy, TB: tuberculosis, IQR: interquartile range, WHO: World Health Organisation, Children: 0-14 years, Adolescents and young adults: 15-24 years, Adults: 25 – 49 years, Elderly: +50 years

The proportion of patients tested for HIV through voluntary counselling and testing among all tested was higher after “Treat All” (37.7% vs 28.6%, p <0.001) while the proportion tested in antenatal care services decreased (11.9% vs 17.6%, p <0.001). There was no significant change in the distribution of patients started on ART by the level of care. The median number of days between HIV diagnosis and ART initiation before and after “Treat All” were 21 (IQR, 5-95) and 1 (IQR, 0-24) days, respectively (S1 Table). The proportion of patients who started ART on the same day increased (43.2% vs 16.4%; p <0.001) (Table 2)

**Table 2:** Comparison of programmatic characteristics before and after the implementation of HIV “Treat All” in 9 pilots districts in Zimbabwe.

| Variables | Categories | Before “Treat All” |  | After “Treat All” |  | <sup>a</sup>p-value |
| --- | --- | --- | --- | --- | --- | --- |
|  |  | n | (%) | n | (%) |  |
| Total |  | 1738 | (100) | 2049 | (100) |  |
| HIV testing modality | Antenatal | 306 | (17.6) | 243 | (11.9) | **<0.001** |
|  | OI, TB & Illness | 580 | (33.4) | 634 | (30.9) |  |
|  | Occupational | 42 | (2.4) | 67 | (3.3) |  |
|  | Voluntary | 497 | (28.6) | 773 | (37.7) |  |
|  | Others | 63 | (3.6) | 52 | (2.5) |  |
|  | Missing | 250 | (14.4) | 280 | (13.7) |  |
| Level of care | Clinic | 1143 | (65.8) | 1337 | (65.3) | 0.421 |
|  | District/Mission Hospital | 460 | (26.5) | 571 | (27.9) |  |
|  | Provincial Hospital | 135 | (7.8) | 141 | (6.9) |  |
| ART Regimen | TDF/3TC/EFV | 1646 | (94.7) | 1962 | (95.8) | 0.221 |
|  | AZT/3TC/NVP(Children) | 43 | (2.5) | 41 | (2.0) |  |
|  | AZT/3TC/LPV/r(Children) | 19 | (1.1) | 17 | (0.8) |  |
|  | Others | 39 | (2,2) | 29 | (1.4) |  |
| Time between HIV diagnosis and ART initiation (days) | Same day | 285 | (16.4) | 885 | (43.2) | **<0.001** |
|  | 1 - 14 days | 342 | (19.7) | 404 | (19.7) |  |
|  | 15 - 90 days | 501 | (28.8) | 283 | (13.8) |  |
|  | > 90 days | 388 | (22.3) | 268 | (13.1) |  |
|  | Missing | 222 | (12.8) | 209 | (10.2) |  |
| <sup>a</sup>Chi-square test, ART: Antiretroviral therapy, OI: opportunistic infection, TB: tuberculosis, TDF: tenofovir, AZT: zidovudine, 3TC: lamivudine, NVP: nevirapine, EFV: efavirenz, LPV/r: lopinavir/ritonavir |  |  |  |  |  |  |

### c) Association between retention and “Treat All”

The maximum follow-up time for the before and after “Treat All” cohorts were 32.9 and 23.8 months while the median was 19.7 (IQR, 6.1 – 28.1) and 16.0 (IQR, 8.9 – 19.7) months respectively. The total attrition for the before “Treat All” cohort was 142/1738 (8,2%; 9.9% stopped ART, 71.8% LTFU and 18.3% died) and after “Treat All” cohort was 165/2049 (8,1%; 23.0% stopped ART, 67.9% LTFU and 9% died) (S1 Table). The proportion of LTFU patients who only visited the health facility once on the day of ART initiation among all patients initiated on ART increased after “Treat All” (2.1% vs 0.5%, p <0.001). Comparing the two cohorts, the 6 and 12 months retention for the before and after “Treat All” were 98.5% (95%CI; 97.8– 99.0) and 95.1% (95%CI; 94.0 – 96.1) and, 97.0% (95%CI; 96.2 – 97.7) and 94.1 % (95%CI; 92.9 – 95.1) respectively (Table 3). Retention during the first 12 months of treatment was higher before “Treat All” (log-rank, p<0.001) (Figure 2).

**Table 3:** Combined, before and after HIV “Treat All” retention in care of patients who started antiretroviral therapy in 9 pilot districts in Zimbabwe.

| Cohort | Months since ART initiation | Retention (%) | 95% Confidence Interval |
| --- | --- | --- | --- |
| Combined | 6 | (97.7) | (97.1 – 98.1) |
|  | 12 | (94.5) | (93.7– 95.2) |
|  | 24 | (88.1) | (86.6 – 89.5) |
| Before “Treat All” | 6 | (98.5) | (97.8– 99.0) |
|  | 12 | (95.1) | (94.0 – 96.1) |
|  | 24 | (90.4) | (88.6 – 92.0) |
| After “Treat All” | 6 | (97.0) | (96.2 – 97.7) |
|  | 12 | (94.1) | (92.9 – 95.1) |
| *After “Treat All”: no patients had a follow-up of 24 months* |  |  |  |

**Table 4.**
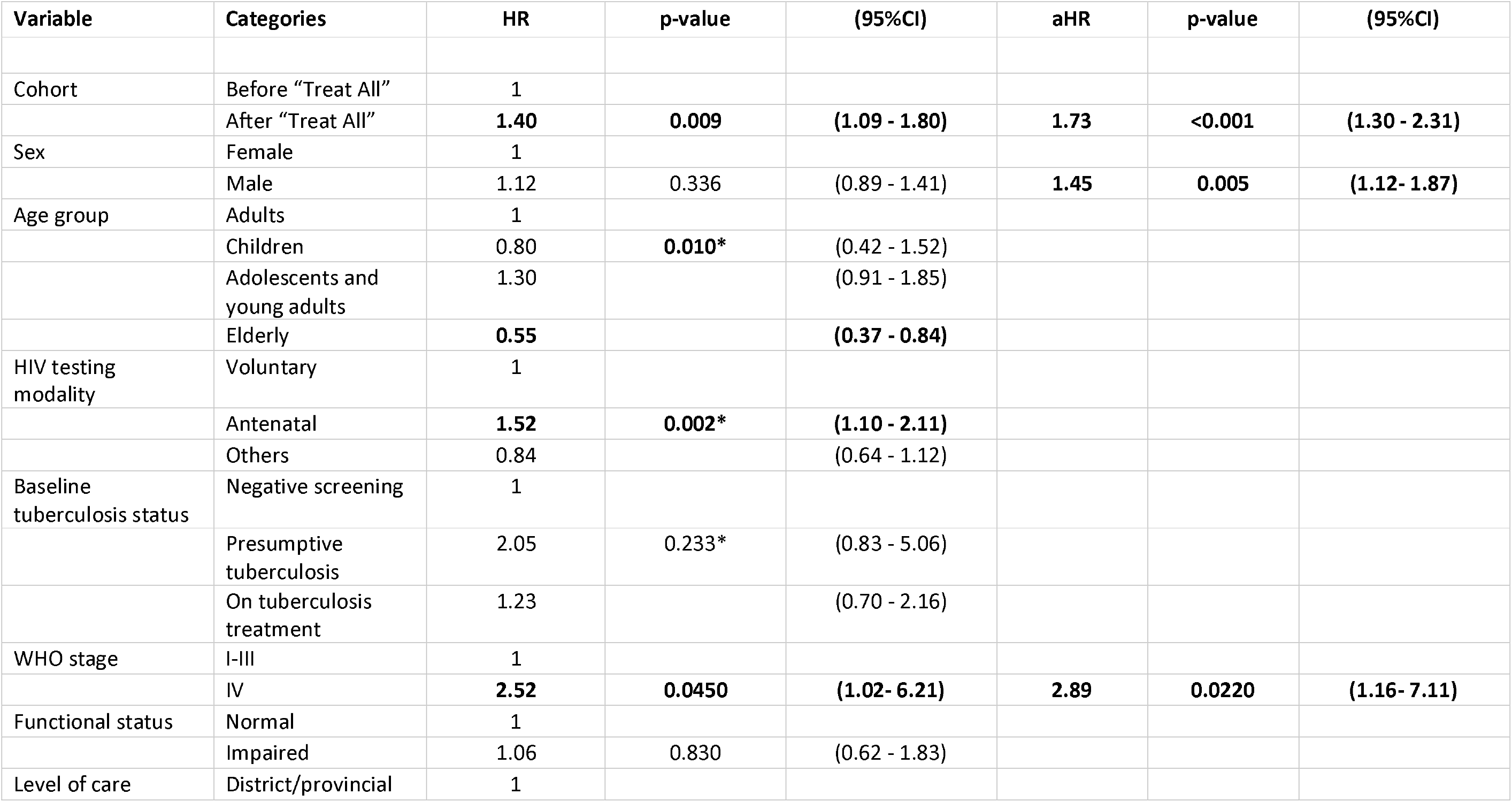
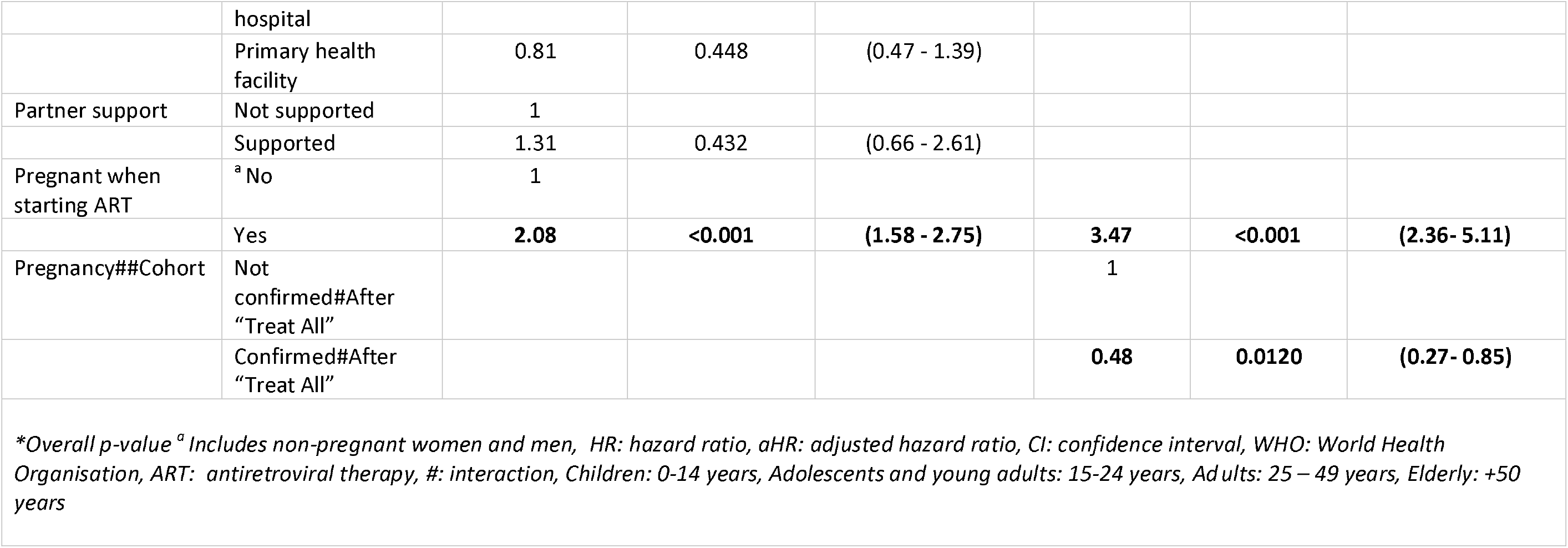
Bivariate and multivariate analysis of factors associated with attrition for patients who started antiretroviral therapy before and after the implementation of HIV “Treat All” in the 9 pilots districts in Zimbabwe (Multiple imputation, N= 3787)

**Figure 1:**
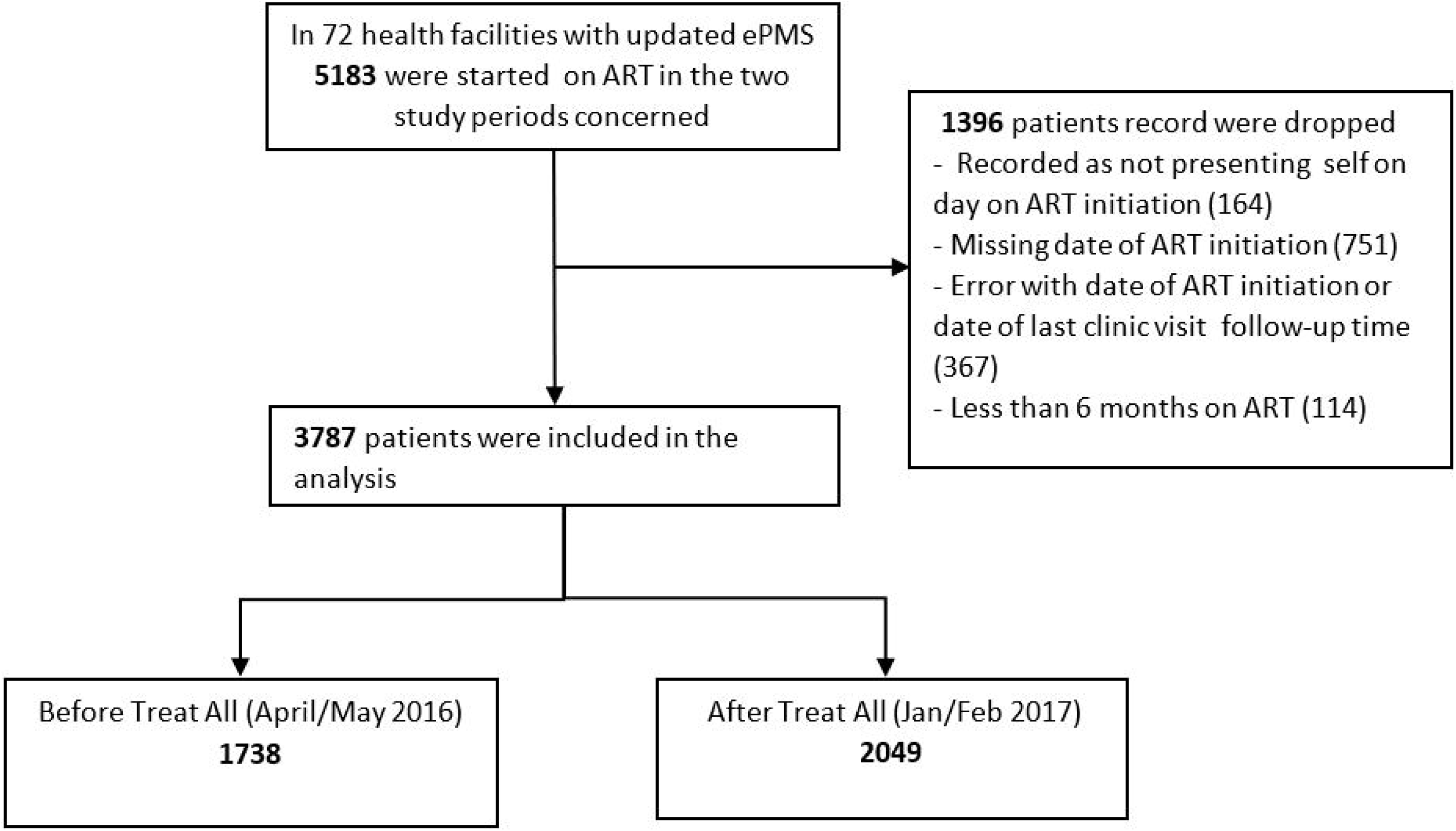
Study participants.

**Figure 2:**
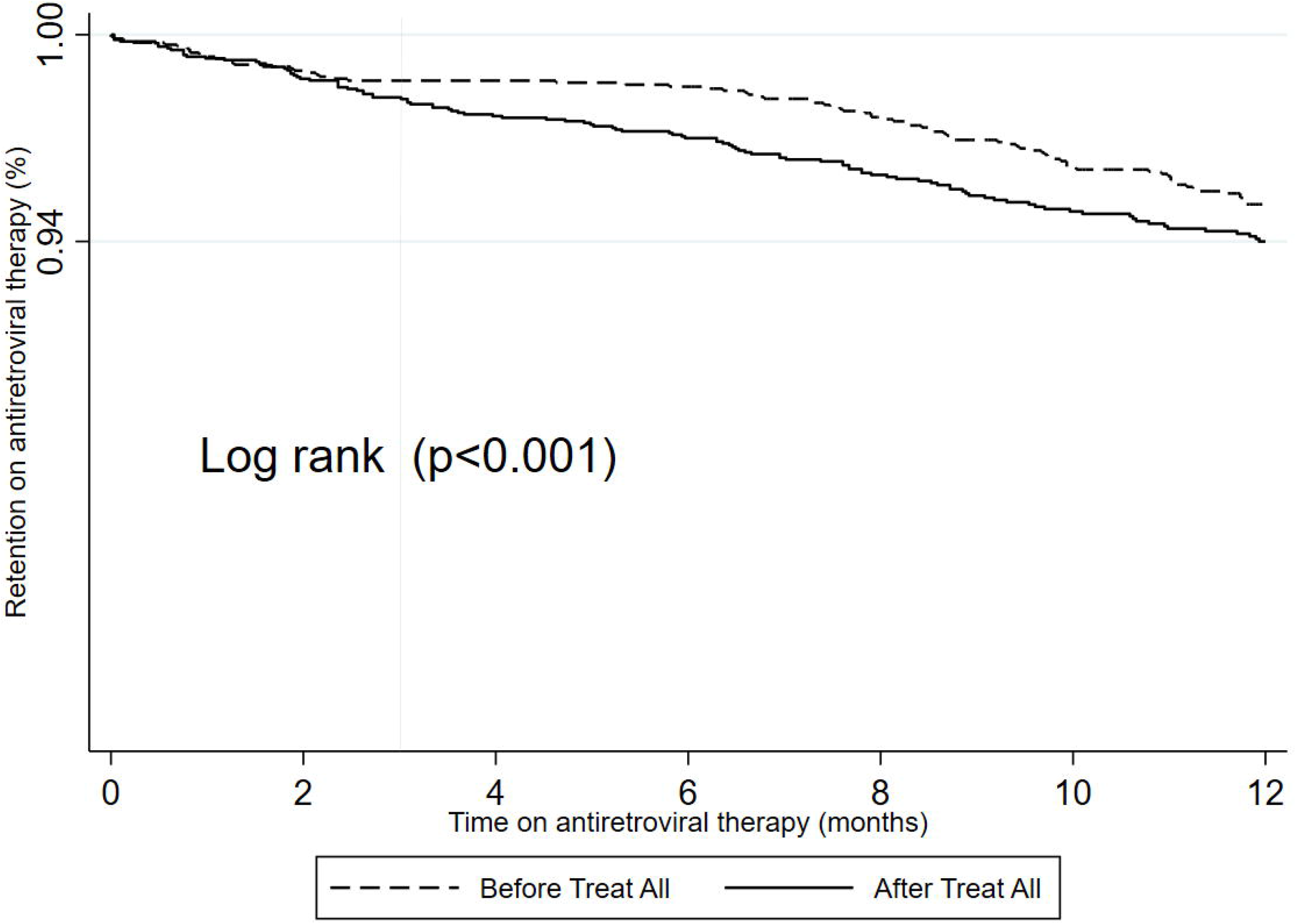
Comparison of the retention at 12 months for patients who started before and after the implementation of HIV “Treat All” in the 9 pilots districts in Zimbabwe.

### d) Predictors of attrition before and after the implementation of “Treat All”

Among non-pregnant women and men, the adjusted hazard ratio (aHR) of attrition after “Treat All” compared to before “Treat All” was 1.73 (95%CI: 1.30 - 2.31). The observed hazard of attrition for women being pregnant at ART initiation decreased by 17% (aHR: 1.73*0.48 = 0.83) since “Treat All”. Being male (vs female; aHR: 1.45; 95%CI: 1.12 - 1.87) and WHO Stage IV (vs WHO Stage I-III; aHR: 2.89; 95%CI: 1.16 - 7.11) predicted attrition both before and after “Treat All” implementation.

The hazard ratio of attrition comparing pregnant women to non-pregnant women was significantly different (aHR: 0.48; 95%CI: 0.27-0.85) after “Treat All” compared to before “Treat All”: the hazard ratio was 3.47 (95%CI: 2.36 - 5.11) before “Treat All” and 1.67 (95%CI:) after “Treat All.”. The association of gender and WHO stage with attrition was not significantly different after “Treat All” compared to before “Treat All”.

## Discussion

### Summary of main findings

This is the first study to assess the performance of the ART programme in Zimbabwe since the scale-up of “Treat All” policy. The study is also a follow-up on previous national studies on ART outcomes in the country. “Treat All” implementation resulted in lower retention and a higher risk of attrition. While the proportion of men among those who started on ART during “Treat All” increased, the proportion of adolescents and young adults among those starting ART did not significantly change between the two periods. After “Treat All” more patients initiated ART on the same day as the HIV diagnosis was made. Being male, WHO Stage 4, and pregnancy predicted attrition in both before and after Treat All. However, pregnancy became a less strong risk factor for attrition after “Treat All” implementation. Our study is among the first to assess if there is a significant change in the predictors associated with attrition before and after the implementation of “Treat All”.

### Association between “Treat All” and patient and programmatic characteristics

In our study, we found the proportion of men in those starting ART under “Treat All” was higher than before. A programmatic report from Lesotho also reported an increase in the proportion of men among those starting ART under “Treat All” [20]. These findings are however contrary to an assessment which was conducted in 6 sub-Saharan African countries to assess the effects of HIV “Treat All” on rapid ART initiation, showing that men remain at risk of not initiating ART [21]. We believe that in our study the reason for a higher proportion of men among those initiating ART might have been due to demand creation, HIV testing and ART initiation strategies that were employed as part of “Treat All” implementation such as moonlight, index case testing and HIV self-testing [22–24]. The increase in patients testing through voluntary HIV testing services, as shown by this study, might also have benefited men.

We found that “Treat All” implementation did not increase the proportion of adolescents and young adults among those starting ART. To date, studies on whether HIV “Treat All” increases rapid ART initiation among adolescents are inconclusive. Positive effects have been shown in some studies, while others have shown that they are still at risk of not initiating ART [21,25–27]. To our knowledge, no evidence-based strategies are currently being implemented in Zimbabwe on a broader scale under “Treat All” to promote ART initiation among adolescents and young adults. Strategies to improve ART initiation among adolescents which include multidisciplinary and adolescent-friendly HIV services together with peer counselling and support, should be tested [28,29]. In our study, we found that more patients are now being initiated on ART on the same day. In clinical trials, same-day ART initiation has shown either no difference or a positive effect on retention [30–33]. Few studies having been done to assess the real-world effects of same-day ART initiation [34], a call for further research. Whether patients are adequately prepared, both psychological and clinically on the same day remains a critical question.

### Association between retention and “Treat All”

To date, the few studies assessing the real-life effects of “Treat All” have shown inconsistent findings regarding its effect on retention. Some have shown patients starting under “Treat All” likely to do better[11,12], others worse [6,7] while others showed no difference [8–10]. In our study, patients who started ART during HIV “Treat All” had lower retention and a higher risk of attrition. This collided with findings from our previous evaluation that patients starting ART at a higher CD4 cut-off might be at risk of attrition [35]. Patients started on ART under “Treat All”, possibly have low-risk perception due to the absence of symptoms and might be less motivated to adhere to lifelong daily ART. [36,37]. The other reason might be during the implementation of “Treat All” early on; more focus has been on ART initiation, including same-day ART initiation. The health care workers were then unable to prepare and provide adequate counselling on the importance of adhering to life-long ART to these asymptomatic patients who might not appreciate its benefits [38]. This is supported by our finding were there was a slight increase of the proportion of LTFU patients who only visited the health facility once on the day of ART initiation among all patients initiated on ART after Treat All. Another study conducted in Eswatini also showed similar findings were the percentage of patients who never returned after the first ART visit doubled (from 3 to 6%) in the “Treat All” period [10]. Several factors can explain the conflicting findings in studies looking at retention during “Treat All”, including different lengths of the follow-up period, settings, methods, study population and definition of outcomes. A systematic review and meta-analysis are needed to synthesise and consolidate the evidence.

The decrease in retention under “Treat All” has also been confirmed by routine programme reports from the national ART programme. This, therefore, calls for the country to develop and implement innovative evidence-based strategies that have been shown to improve retention. The process has already started with differentiated ART delivery models (Community ART refills groups-CARGS, adherence clubs, fast track and outreach) currently being scaled-up. By the end of 2018, around 30% of stable adolescents and adults on ART were receiving ART through at least one differentiated ART delivery model [39]. Systems to track and trace defaulters have also been put in place. Despite systems to track defaulters, LTFU continues to be a challenge. LTFU accounts for most of the attrition. This is thought to have also an administrative reason [35], and the MoHCC and the supporting implementing partner have since instituted a National ART Census Survey to be able to get correct estimates of the number of patients on ART. The results are yet to become available in the public domain.

### Predictors of attrition before and after the implementation of “Treat All”

In our study pregnancy became a less strong risk factor for attrition since “Treat All” implementation, which is a positive development. We hypothesise that the reduction might be due to the expansion of “Treat All” initiative to the rest of PLHIV. For a long period, “Treat All” has been specifically focused on the subgroup of HIV positive pregnant women and delivered as Option B+, which might have created some “stigma” [40,41]. Now, this is applied to all patients; this label might have been removed. Current efforts to retain pregnant and breastfeeding women on ART which include the integration of services, family-centred approaches, and the use of lay healthcare providers which include mentor mothers should be optimised and continued [42,43]. Documentation and follow-up of HIV positive pregnant women with complications referred from lower to higher-level health facilities (maternal waiting homes) should also be improved so that these referrals are not treated as LTFU.

In the current study, adolescents and young adults had comparable retention with adults contrary to other studies. The finding is contrary to other programmatic HIV “Treat All” studies to date [6,7,9–11]. Zimbabwe has employed strategies to retain adolescents in care, mainly through its home-grown Community adolescent’s treatment supporters (CATS) model. Evidence has shown the CATS model to be effective in retaining adolescents in HIV care [44]. To date, the model has been scaled up to most of the districts. Resources to fully implement the CATS model throughout the country should be mobilised.

We found men to be at risk of attrition under “Treat All”, a finding similar to other studies [7,9]. Differentiated service delivery strategies that have been shown to retain men in ART care under HIV “Treat All” which includes tailored awareness on the health benefits of early ART start, accelerated linkage to care, decreasing logistical barriers to HIV care, patient-centred approach, flexible clinic hours and tracking and tracing of those missing appointments should be strengthened, maintained and optimised [45].

### Strength and limitations

Compared to other studies, our study sample size was large and representative. The pilot districts were distributed across all the regions of the country. The study was the first to assess the performance of the ART programme since the start of HIV “Treat All” policy implementation and allowed us to test and confirm findings from our previous evaluation.

However, our study had limitations. We evaluated the effect of HIV “Treat All”, considering it as one intervention, though it has several components across the cascade of care. We were unable to disentangle these components and assess their individual effects on attrition. Our study was based on health facilities with ePMS, which is only available in high volume health facilities, and this might affect the generalisability. However, sites with ePMS contain most patients on ART in the country. We could also not assess re-engagement in care since the ePMS database only reports the most recent status of the patient with no record of previous disengagements. Because we used routine electronic programme data collected retrospectively, we had to deal with missing data. Data on weight, height, CD4 and viral load were more than 50% missing, so could not be assessed as a confounder. To assess the effect of disease progression on attrition, we then used WHO Staging and functional status as proxies in our model. Most of the attrition in our study was explained by LTFU. Some of these patients reported as LTFU might be still alive and on ART (unreported self-transfers), stopped ART or unreported deaths [46].

Because of missing data, we were also unable to compare the viral load suppression rates between the before and after HIV “Treat All” cohorts. Unfortunately, it was impossible to solve this by merging ePMS and laboratory data, due to missing or inconsistent format of the unique patient identifier between the various testing laboratories. Strategies to improve data quality and making sure the existing electronic data collection systems are interoperable need urgent attention. This is an observational study looking into an association of individual patient and programme characteristics and retention in care. We are aware that lumping patients from all facilities together may have caused a loss of information on specific health facility-level factors that influence retention, and that could not be studied.

### Future research

Considering inconsistency of evidence in the programmatic effects of “Treat All”, there is a need for more research in the domain. From our field experience, we have noticed the implementation of “Treat All” encompasses many aspects across the cascade of care. The package includes innovative HIV testing and linkage to care strategies, clinical and psychological readiness assessment, retesting before ART initiation, same-day ART initiation, follow-up and retention in care strategies. On top of the HIV cascade of care issues; there are also health systems challenges anticipated from the abrupt increase in the number of patients now being followed-up in care. Further research, both quantitative and qualitative, should explore the influence of all these factors on “Treat All” implementation to optimise it. Men continue to be at risk under “Treat All”; further research is needed on how ART delivery can be streamlined to retain them in care. Most studies have focused on patient-level factors on ART outcomes. This, therefore, calls for studies looking into the impact of health facility-level characteristics on ART outcomes since performance might vary across facilities. Finally, as more studies looking into the real-world effects of “Treat All” surface, there is a need for systematic reviews and meta-analyses summarising outcomes across the cascade of care and evaluation of strategies to optimise “Treat All” implementation.

## Conclusion

In conclusion, our study found that since the implementation of “Treat All” the retention in care was lower and the risk of attrition was higher. Male sex, advanced HIV disease and pregnancy are risk factors for attrition both before and during the implementation of HIV “Treat All”. However, after the implementation of “Treat All” pregnancy became a less strong risk factor. Strategies to retain men and to improve ART initiation among adolescents and young adults should be prioritised. Patients with advanced disease (WHO Stage IV) continued to be at risk of attrition. Further research should explore how different components of HIV “Treat All” implementation (HIV testing, linkage, same-day ART initiation, clinical and psychological preparation) and health system/facility issues have a bearing on patient outcomes.

## Data Availability

The study was conducted with routinely collected program data of the Zimbabwe National ART Programme available in an electronic patient management system (ePMS) However, the data is not available on the public domain, and anyone interested in using the data for scientific or academic purpose should contact the Director of the AIDS and TB Program, Ministry of Health and Child Care, Government of Zimbabwe, 2nd Floor, Mukwati Building, Harare, Zimbabwe.

## Acknowledgements

The authors would like to thank the following organisations for support during the study - Ministry of Health and Child Care (MoHCC); the University of Zimbabwe, College of Health Sciences (UZCHS), Department of Community Medicine; Institute of Tropical Medicine (ITM) in Antwerp, Belgium; the University of Zimbabwe, Zimbabwe AIDS Prevention Project (ZAPP-UZ) and PEPFAR Zimbabwe and its main implementing partners, I-TECH and OPHID for supporting the HIV “Treat All” pilot. The authors are grateful to the team that assisted in ePMS data management prior to analysis. We also want to various stakeholders and implementing partners who have been supporting the MoHCC towards improving the MoHCC AIDS and TB Program data quality through various initiatives, including supporting the ePMS. Of note is the Global Fund for AIDS, Tuberculosis, and Malaria (GFATM) which supported MoHCC in setting up the ePMS. Finally, we thank front line health workers and PLHIV on ART whose records were used.

## Authors Contributions

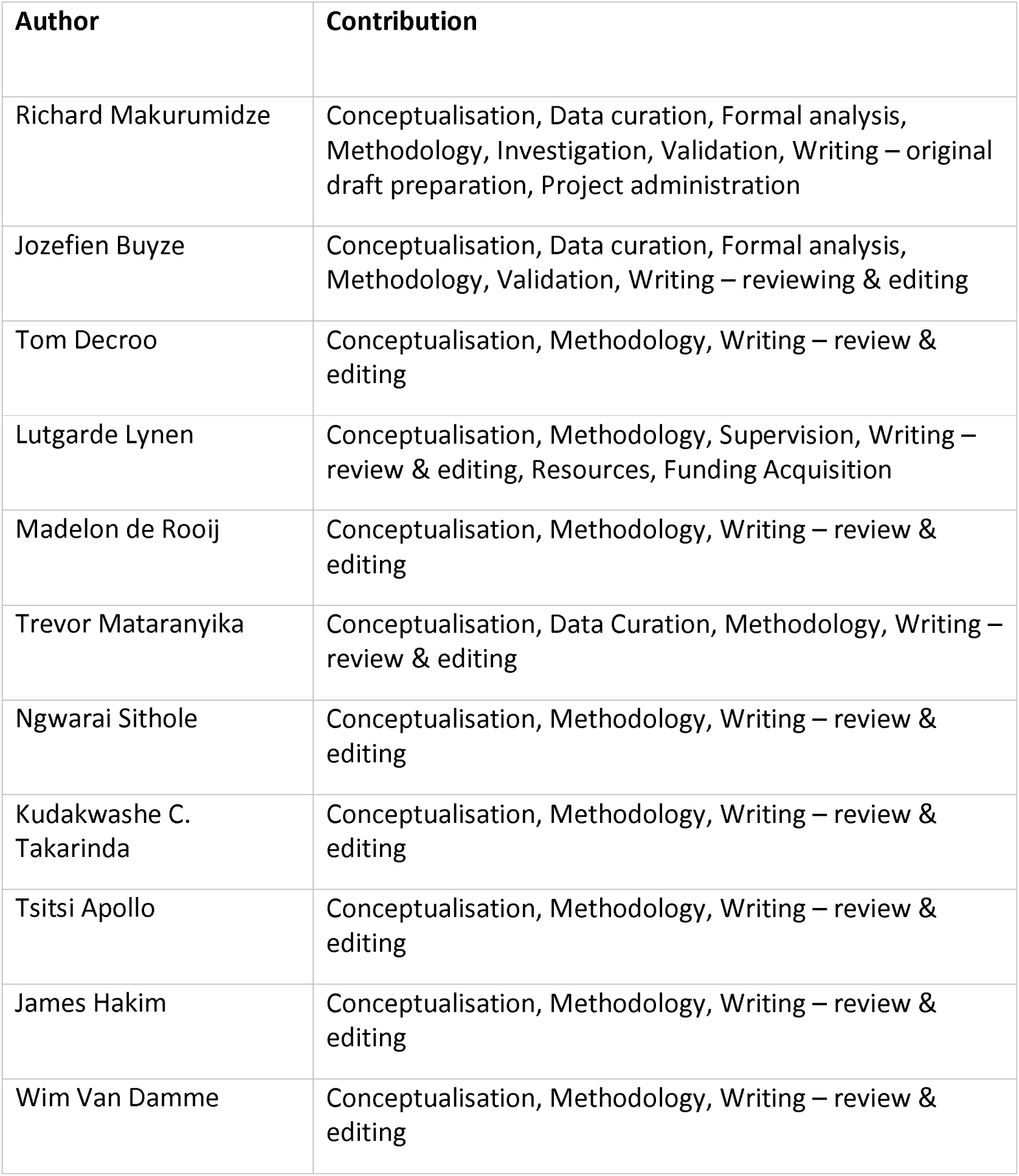
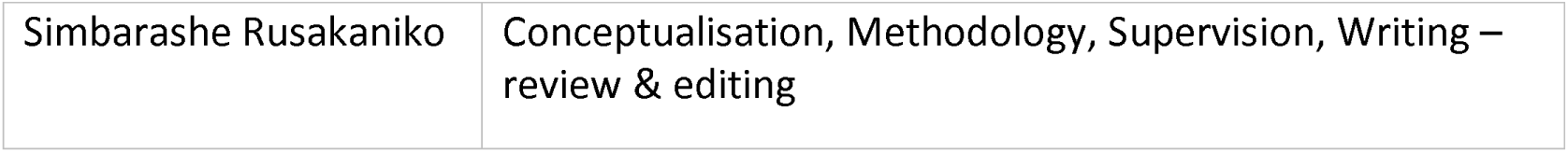

## Declarations

### Funding

Richard Makurumidze receives a PhD scholarship grant from the Institute of Tropical Medicine, funded by the Belgian Development Cooperation (DGD). The funders had no role in study design, data collection and analysis, decision to publish, or preparation of the manuscript.

### Competing Interests

The authors declare that they have no competing interests.

## Supporting information

**S1 Figure:** Data management system for the National ART Program in Zimbabwe

**S1 Table:** Outcomes, follow time and time from testing to ART initiation for patients who started antiretroviral therapy before and after the implementation of HIV “Treat All” in the 9 pilots districts in Zimbabwe.

**S2 Table:** Attrition for patients who started antiretroviral therapy before and after the implementation of HIV “Treat All” in the 9 pilots districts in Zimbabwe

**S1 File:** Retention and predictors of attrition among patients who started antiretroviral therapy in Zimbabwe’s national antiretroviral therapy programme between 2012 and 2015. PLoS ONE 15(1): e0222309. https://doi.org/10.1371/journal.pone.022230

## Notes

### Competing Interest Statement

The authors have declared no competing interest.

